# Peripheral innate and adaptive immune cells during COVID-19: *Functional neutrophils, pro-inflammatory monocytes and half-dead lymphocytes*

**DOI:** 10.1101/2020.08.01.20166587

**Authors:** Emel Ekşioğlu-Demiralp, Servet Alan, Uluhan Sili, Dilek Bakan, İlhan Ocak, Rayfe Yürekli, Nadir Alpay, Serpil Görçin, Alaattin Yıldız

## Abstract

A better understanding of the innate and adaptive cells in the COVID-19 disease caused by the SARS-CoV-2 coronavirus is a necessity for the development of effective treatment methods and vaccines. We studied phenotypic features of innate and adaptive immune cells, oxidative burst, phagocytosis and apoptosis. One hundred and three patients with COVID-19 grouped according to their clinical features as mild (35%), moderate (40.8%), and severe (24.3%) were included in the study. Monocytes from all COVID-19 patients were CD16^+^ pro-inflammatory monocytes. Neutrophils were mature and functional. No defect has been found in ROS production of monocytes and neutrophils as well as no defect in their apoptosis. As bridging cells of the innate and adaptive immune system; the percentage of NK cells was in normal range whereas the percentages of CD3^-^CD8^+^CD56^+^ innate lymphoid and CD3^+^CD56^+^ NK like T cells were found to be high in the severe cases of COVID-19. Although absolute numbers of all lymphocyte subsets were low and showed a tendency for a gradual decrease in accord with the disease progression, in all COVID-19 patients, the lymphocyte subset with the most decreased absolute number was B lymphocytes, followed by CD4 + T cells in the severe cases. The percentages of suppressive, CD3^+^CD4^-^CD8^-^; HLA-DR^+^CD3^+^ and CD28^-^CD8^+^ cells were found to be significantly increased. Importantly, we demonstrated spontaneous caspase-3 activation and increased lymphocyte apoptosis. Altogether our data suggest that SARS-CoV-2 primarily affects lymphocytes not innate cells. So that, it may interrupt the cross-talk between adaptive and innate immune systems.

## INTRODUCTION

The SARS-CoV-2 virus, originating from Wuhan, China since December 2019, causing a worldwide pandemic is a positive-sense single-stranded RNA virus (1, 2). The disease caused was called Coronavirus disease 2019 (COVID-19). Although the disease predominantly causes pneumonia, the first identified route of uptake into the cell is through ACE-2 receptors (3, 4). Other organ involvement is possible since the ACE-2 receptor is commonly found in the body(5).

Indeed, as clinical experience has accumulated, it has been shown to affect other organs and tissues in COVID-19 disease(6). Although the Coronavirus family to which SARS-CoV-2 belongs has similarities with the Middle East Respiratory Syndrome (MERS) and Severe Acute Respiratory Syndrome (SARS) viruses, it has also been shown to have higher virulence and stronger attachment to the ACE-2 receptor(4).COVID-19 disease can progress mildly (81%), moderate to severe (14%) according to pulmonary involvement, and critical (5%) with multiple organ failure(7). The recent mortality rate has reached 6.3%, although it varies with age, gender, occupational group, and the presence of comorbidities such as diabetes, hypertension (8–10).

First-line defense against intracellular invaders such as viruses starts with innate immune cells including monocytes, macrophages, neutrophils. Activation of monocytes, macrophages, and bystanders, bridging dendritic cells by pattern recognition receptors results in releasing of proinflammatory cytokines such as IL-1, IL-18, IL-33, and IL-6. Those cytokines activate Natural Killer (NK) cells as well as producing a positive feedback loop for innate cells and also for adaptive immune cells. NK activation is critical for viral defense because of both their cellular cytotoxic function as well as their cytokine release in particular interferon-gamma that activates both B and T cells(11). On the other hand, adaptive activation is a requirement for both eliminating invaders, constructing a proper immune memory, and also for limiting innate activation. Studies in T cell and RAG-1 deficient mice demonstrated that lack of lymphocytes resulted in cytokine storm syndrome and death (12). Crosstalk between natural and adaptive immune systems constitutes our entire immune system, but every fine-tuned interaction is a target point for invaders.

The vicious circle of cytokine storm syndrome and hyperinflammation in COVID-19 deteriorate patients rapidly and mortality rate increases in this group due to respiratory failure (13). One of the main reasons for the rapid deterioration of COVID-19 patients is hyper inflammation and cytokine storm syndrome. Both start with increased interleukin 6 (IL-6) and IL-1B by activation of the NLRP3 inflammasome(14, 15). Although the viral protein that activates the NLRP3 inflammasome in SARS-COV2 is unknown, the SARS-COV virus is known to activate the NLRP3 inflammasome through the viroporin 3 protein and causes high levels of IL-1B release (15). Studies show that SARS-COV2, like other coronaviruses, use only the NfKB signaling pathway that activates innate cytokines such as IL-1, IL-6, and TNF-a. More, it uses strategies to inhibit the interferon signaling pathway (16).

Although what is known about immune pathogenesis of the disease is very limited, lymphopenia, the change of neutrophil/lymphocyte ratio in favor of neutrophils, increased level of LDH, CRP and ferritin vary in proportion with the severity of the disease and are used in the follow-up of the disease (17–19). In the autopsies of the individuals who died due to Covid-19, neutrophil, and monocyte/macrophage infiltration and few helper T cells were detected in their lungs (20, 21). It is noteworthy that the natural killer (NK) and CD8^+^ cytotoxic T cells, which are of primary importance in the fight against viral infection, are not present in the field (22). Considering lymphopenia, it brings the suspicion that the SARS-CoV-2 virus may attach other surface and cytoplasmic molecules other than ACE-2, and may also target the lymphoid system apoptosis similar to SARS-CoV virus 7a protein which induces apoptosis by competing to bind anti-apoptotic member of Bcl-2 family, Bcl-XL (23). Immune profiles of individuals with Covid-19 disease should be examined and be informed more to establish explanatory immune hypotheses on this disease. Indeed, it has been shown that a low number of CD4^+^, CD8^+^ T lymphocytes, NK cells as well as B lymphocytes (24, 25). Functions of T lymphocytes measured by interferon-γ release following stimulation has been found to be in the normal range (25). In this cross-sectional study, we aimed to examine and compare peripheral innate and adaptive cells, burst and phagocytosis functions of monocyte and neutrophils and lymphocyte apoptosis of patients with COVID-19 disease in mild, moderate and severe disease courses. The data compared against the normal values of the individuals used as a healthy control group in previous studies of the group or the defined normal values of the subgroups. Delineation of the phenotypic and functional impairments created by the SARS-CoV-2 virus in the cells of the innate and adaptive immune systems will help to better recognize the virus and develop treatment and prevention strategies.

## PATIENTS and METHODS

### Patients

A total of 103 patients infected with the new Coronavirus-2019, who applied to Istanbul Memorial Şişli Hospital and Marmara University Pendik Training and Research Hospital with various symptoms and signs from sore throat to shortness of breath were included in the study [52 female, 51 male with the mean age of 53,9 ± 15 (25-88)]. Covid-19 RT-PCR positivity was the inclusion criterion in the study (tested by Covid-19 kit, Bioeksen R&D Istanbul, Turkey). However, a group of patients was diagnosed by computed tomography (CT) with the ground glass appearance in their lungs. Thus, 76.7% of our patient group was PCR and CT positive; 16.5% of the patients were only PCR positive, outpatient group; and 6.8% were only CT positive. 52.4% of our patient group also had chronic diseases such as diabetes, hypertension, coronary artery disease, and renal insufficiency. This subgroup was also separately evaluated in immunological analyses. Mean hospitalization time was 8 ±8.5 days (range:1-51; Table 1).

**TABLE 1.** Patients’ Characteristics.

| Gender | | Age | SARS-CoV2 + | | | Presence of | Mean duration ±SD |
| --- | --- | --- | --- | --- | --- | --- | --- |
| Female | Male | (range) | PCR | CT | PCR and CT | Chronic Diseases | (range) |
| 52 | 51 | $53,9 \pm 15$ | 16.5% | 6.8% | 76.7% | 52.4% | $8 \pm 8.5$ days |
| Total:103 |  | (25-88) |  |  |  |  | (1-51) |

The patients were clinically classified; a group of patients with mild CT findings without lymphopenia (percentage>20%) were added to the outpatient group and classified as “mild group (35%)”; patients who had borderline lymphopenia (percentage: between 20% −15%) plus more common CT involvement, but with oxygen saturation above 90%, were classified as ‘moderate group (40.8%)’; in addition to widespread lung involvement, lymphopenia (percentage<15%) and patients with oxygen saturation below 90% were grouped as ‘severe group (24.3%)’ and their immunological features were examined accordingly. According to the instructions of our Ministry of Health for patient treatments; That Hydroxychloroquine alone or hydroxychloroquine and azithromycin combination in mild patients; favipiravir in addition to the combination of hydroxychloroquine azithromycin in moderate patients; additional favipiravir and/or tocilizumab and/or convalescent plasma treatments in addition to the hydroxychloroquine azithromycin immunological parameters of our patients according to their treatment strategies in this cross-sectional study.

The study was approved by both Ministry of Health, Scientific Research Platform (no:2020-04-30_11_31) and Marmara University Ethics Committee (no:08.05.2020/09.2020.541). Informed consent was obtained from the participants before the study.

### Controls

In the comparison of immunological parameters with healthy controls, raw flow cytometric data from healthy controls that have been tested before for the laboratory routine norm studies in Ekşioğlu-Demiralp’s Laboratory and samples that have been used as controls to the diseases in the theses instructed by Ekşioğlu-Demiralp were used. The gender and age distributions of the controls were selected in accordance with the Covid-19 patient group. A total of 100 healthy controls have been included in comparisons.

### Isolation of White Blood Cells and Immunophenotyping

Peripheral blood White Blood Cells (WBC) of all participants were isolated from their hemogram tubes with EDTA taken for their routine tests by using erythrocytes lysing solution (155 mM NH4Cl; 10 mM KHCO3; 0,1mM EDTA; pH:7.3). The following fluorochrome labelled monoclonal antibodies (mAb) and isotype-matched controls were used for two-three color phenotypic analysis: anti-IgG1, anti-IgG2a, anti-CD45, anti-CD3; anti-CD4; anti-CD8; anti-CD16; anti-CD56; anti-CD19; anti-CD20, anti-HLA-DR; anti-CD10; anti-CD25; anti-CD28, anti-CD69, (Becton&Dickinson Inc, San Jose, CA, USA). Cells were acquired and analyzed using CellQuest software on a FACSCalibur flow cytometer (Becton Dickinson Inc, San Jose, CA, USA). Lymphocytes, monocytes, and neutrophils were gated according to their forward and side scatter characteristics and their specific CD markers. Populations were evaluated as percentage. The absolute number of subsets was calculated in accordance with the absolute number of lymphocytes for each individual sample. Multiple gating strategies were applied when required. Mean fluorescence intensities (MFIs) of CD10 and CD16 antigens on neutrophils and MFIs of CD16 and HLA-DR on monocytes were also evaluated as an indication of activation/exhaustion criterion.

### Oxidative Burst and Phagocytosis

White blood cells from 13 patients in moderate clinical course and 5 healthy controls were isolated from their heparinized peripheral blood samples and simultaneously tested for oxidative burst and phagocytosis functions of their neutrophils and monocytes by applying instructions of a standard kit (PhagoBurst Test, Becton Dickinson Inc, San Jose, CA, USA). Percentage results were evaluated by using the test’s standard values and, also by comparing to controls. Moreover, the mean fluorescence intensity (MFI) of each PMA-stimulated sample was divided into that of its non-stimulated sample. Thus, the folds increases in DHR123 fluorescence following PMA stimulation were calculated for neutrophils and monocytes and compared between patients with COVID-19 and the controls.

### Apoptosis and Cell Death

White blood cells from 13 patients in moderate clinical course and 5 healthy controls were isolated from their heparinized peripheral blood samples. A cell-permeable fluorogenic caspase-3 substrate, Phi-Philux.G1D2 (OncoImmunin Inc, Kensington, MD), and propidium iodide (PI) were used to evaluate early, late apoptosis and cell death. Briefly, cells (1×10^5^/ml) were resuspended in phosphate-buffered saline (PBS) containing 5 % fetal bovine serum (FBS) and one tube was stimulated for apoptosis using 100 ng/ml of PMA at 37^0^C for 1 hour, while other was incubated at the same temperature without stimulation, as a control. Then 9 mM Phi-Philux.G1D2 was added to the tubes and samples were incubated for an additional one hour at the same conditions. Following the incubation, PI (20mg/ml in PBS) was added to evaluate cell death simultaneously. Then the cells were immediately acquired by flow cytometry (FACSCalibur, Becton Dickinson Inc, San Jose, CA, USA) using CellQuest software, where at least 40,000 cells were acquired. PhiPhilux stained only cells were evaluated as early apoptosis or activation; PhiPhilux+PI stained cells as late apoptosis; and PI stained only cells as cell death. All were assessed in lymphocyte, monocyte, and neutrophil gates. Initial MFI values of PhiPhilux fluorescence in unstimulated samples that demonstrate the spontaneous activation status of the cells were measured and compared.

### Statistical analysis

The statistical software package SPSS version 25.0 was used for all statistical analyses. Values were presented as percentages and absolute numbers ± standard deviation of the mean. Statistical significance of differences between the groups was calculated using with an analysis of variance (ANOVA) and followed by Bonferonni test as post-hoc. The significance of differences in pre-after measurements was calculated using the Wilcoxon signed ranks test. Mann-Whitney U test was used for non-parametric comparisons in small groups. Values of p<0.05 were regarded as significant.

## RESULTS

### Leucocyte counts and percentages

WBC counts, and percentages of lymphocytes, monocytes, and neutrophils and their absolute numbers in patients with COVID-19 were summarized in Table 3. Accordingly, there was no difference in total WBC numbers between the groups. The lymphocyte percentage was found to be significantly lower in the moderate and severe groups compared to the mild group (p<0.01, Table 3). Absolute lymphocyte count was lower in the moderate and severe group compared to the mild group, but it reached statistical significance only between the mild and severe group (p=0.02, Table 3). The percentage of monocytes was higher in mild COVID-19 patients compared to moderate and severe patients (p<0.05). However, there was no statistically significant difference between the groups in absolute monocyte counts. The percentage of neutrophils percentage was low in the mild group. Statistical difference was found only between mild and severe groups (p <0.01, Table 3). In addition to the increase in percentages of neutrophils, absolute number of neutrophils was also found to be statistically high in the moderate and severe groups in comparison to the mild group (p <0.01, Table 3).

**TABLE 2.** Patients’ Clinical Characteristics.

|  | n (%) | PCR | CT | O<sub>2</sub> Saturation | Lymphocytes (%) | Treatment* |
| --- | --- | --- | --- | --- | --- | --- |
| **MILD** | 36 (35%) | + | -/+ | >90% | >20 % | HQ or HQ+A |
| **MODERATE** | 42 (40.8%) | +/- | + | >90% | 20%>L>15% | HQ+A + F |
| **SEVERE** | 25 (24.3%) | + | + | <90% | <15% | HQ+A+F + Tc and/or P |

\*: HQ: Hydroxychloroquine; A: Azithromycin; F: Favipiravir; Tc: Tocilizumab; P: Plasma treatment

**TABLE 3.** Patients’ leucocyte number and percentages in various COVID-19 disease course.

|  | **WBC**<br>(cells/mm<sup>3</sup>) | **Lymphocytes**<br>(%) /(cells/ml) | **Monocytes**<br>(%) /(cells/ml) | **Neutrophil**<br>(%) /(cells/ml) |
| --- | --- | --- | --- | --- |
| **MILD** | 5.4±2 | 30±11.8* (1486±827)# | 10.9±3.9* (532±250) | 57.3 ± 14* (2988±1690)# |
| **MODERATE** | 6.1±3.3 | 19.2±10.8 (1060±929) | 8.1±3.8 (449±241) | 70.8 ± 13 (4485±2981) |
| **SEVERE** | 5.8±2.7 | 15.1±8.9 (876±676)# | 9.1±4 (481±333) | 74.5 ± 10.9 (4300±2081) |
| **p value** | NS | *: $p < 0.01$; # $p = 0.02$ | *: $p < 0.05$; NS | *: $p < 0.01$; # $p < 0.01$ |

Thus, it has been shown that lymphopenia deepened and granulocytosis increased, as both percentage and absolute number proportionally to the severity of the disease in accordance with the literature (19). There was no difference in either WBC counts or percentages of leukocyte subsets when patients were classified according to their co-morbidities.

### Phenotypic and functional features of innate immune system cells in COVID-19

Phenotypic features of monocytes, neutrophils, NK and NK-like innate lymphoid cells and, oxidative burst, phagocytosis and apoptosis of monocyte and neutrophils were assessed.

### Pro-inflammatory monocytes with decreased HLA-DR expression in patients with COVID-19

As antigen-presenting cells, monocytes constitutively express HLA-DR. In our study, the mean percentage of HLA-DR expression on monocytes was 86.2%±10.4 in the mild; 85.7%±10.7 in the moderate and 85.3%±8.8 in severe groups of patients. That was 94.5 %±3.6 in the healthy control group and HLA-DR expression on monocytes of all three groups was significantly lower in comparison to healthy controls (p<0.01; Fig 1a). Although there was no difference in the percentages of HLA-DR among the patients’ groups, Mean Fluorescence Intensity (MFI) of HLA-DR, which is a relative indicator of the number of molecules per unit cell, was 521 ± 293 in the mild; 299 ± 200 in the moderate and 294 ± 192 in the severe group of patients with COVID-19 (Fig 1b). Mild group MFI values were significantly higher in comparison to that of moderate and severe patients (p=0.02, p=0.05 respectively). HLA-DR MFI on monocytes of the healthy population was 752 ± 352 significantly different from all patients group (p=0.02 for mild, p<0.01 for both moderate and severe patients; Fig 1b). The gradual decrease of HLA-DR expression on monocytes of patients with COVID-19 can be considered as a gradual exhaustion of monocytes while disease progresses. The expression of CD16 defines a subgroup of monocytes which is called intermediate, non-classical monocytes. It has been shown before that they were pro-inflammatory monocytes (42). We demonstrated that monocytes of all groups of COVID-19 expressed CD16 whereas that expression was very low in the healthy control group (69%±23 in the mild; 57.7% ± 27 in the moderate; 52.6% ± 32 in the severe group of patients versus 8.7%±6 in controls, p<0.01 Fig 1c, d). That difference was also significant between the mild and severe cases (p<0.05), Monocytes from patients with COVID-19 tended to lose their CD16 during disease progression and that the mean percentage of CD16 on monocytes was negatively correlated with the clinical course (CC: −0.24; p = 0.03) has been shown. In the severe cases, that decrease might be a second proof of that monocytes have been exhausted, similar to losing their HLA-DR on their surfaces when the disease becomes serious.

**FIGURE 1.**
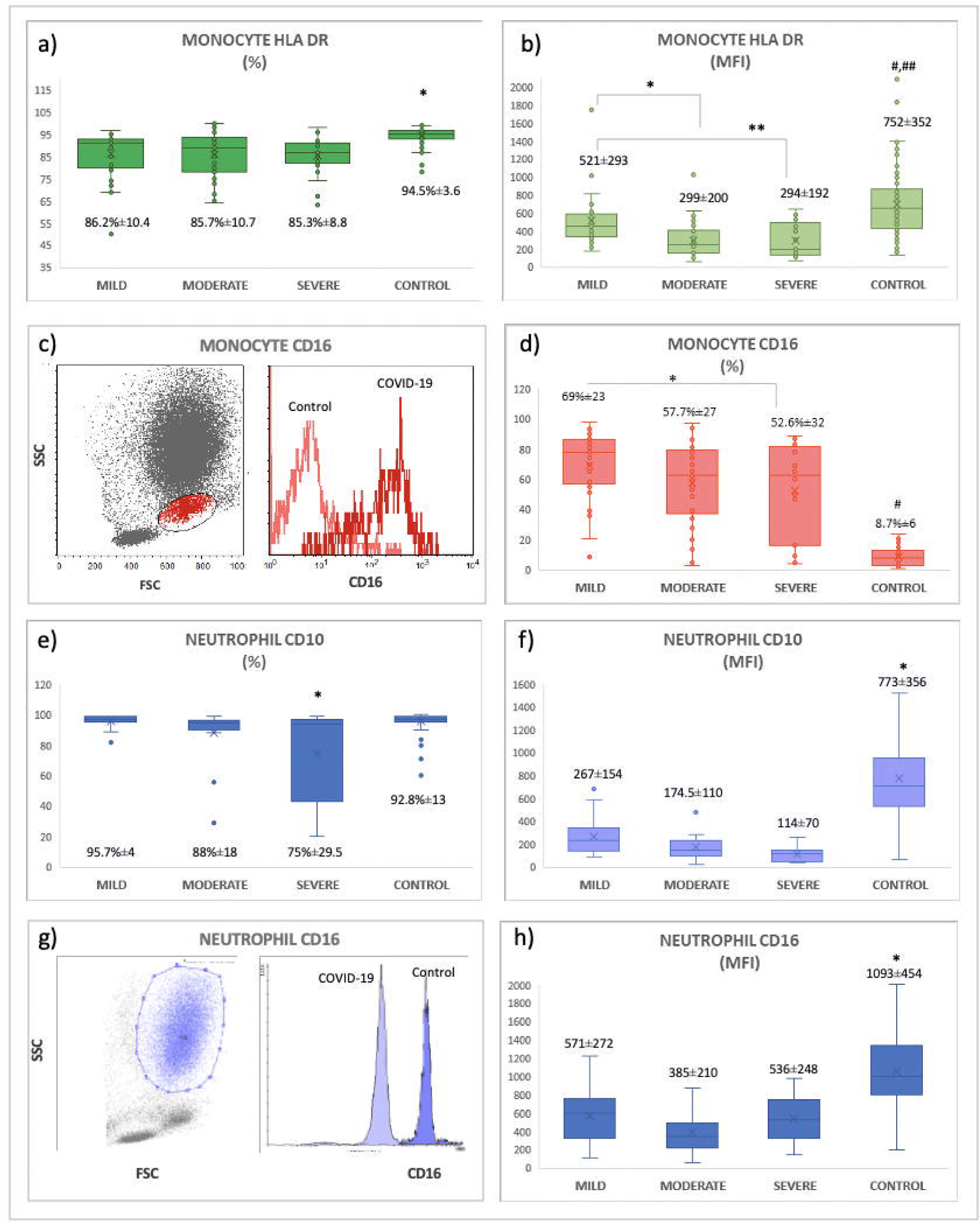
Comparisons of phenotypic features of monocytes and neutrophils in patients with COVID-19 and controls. **a)** HLA-DR expression on monocytes from Covid-19 patients from all clinical courses is significantly low compared to controls. *: p<0.01 **b)** MFI values of HLA-DR on monocytes from Covid-19 patients with the mild clinical course is significantly different from the patients with moderate and severe disease course (*: p=0.02; **: p=0.05 respectively). All patients’ groups have decreased HLA-DR MFI compared to the control group (#: p=0.02 for mild patients; ##: p<0.01 for moderate and severe patients). **c)** A representative plot showing gate for monocytes on FSC vs SSC and an overlaid histogram demonstrating CD16 expression on monocytes from a patient with COVID 19 and healthy control. **d)** The expression of CD16 on monocytes gradually decreases depending on the disease course (*:p=0.05 between the mild and severe groups). But, all were significantly high in comparison to controls (#: p<0.01). **e)** The expression of CD10 on neutrophils is significantly low in COVID-19 patients with the severe course (*: p<0.02). **f)** MFI values of CD10 on neutrophils in patients with COVID-19 are low compared to controls. (*: p<0.01) **g)** A representative plot showing gate for neutrophils on FSC vs SSC and an histogram demonstrating CD16 expression on neutrophils from a patient with COVID 19 and healthy control. **h)** CD16 MFI values of neutrophils from patients with COVID-19 with the different clinical course were decreased in comparison to controls (*: p<0.01)

### Expressions of CD10 and CD16 on neutrophils from COVID-19 patients

As the markers of mature neutrophils, expression of CD10, and CD16 were measured on neutrophils. That CD10 expression 95.7%± 4 in the mild; 88%± 18 in the moderate; 74.9%± 29.5 in the severe group patients with COVID-19, and 92.8%± 13 in the controls have been found. CD10 expression was found to be significantly low in the severe group compared to the others (p<0.02; Fig 1e). When MFI of CD10 was measured; a consistent decrease has been found in all three groups (267±154 in mild patients, 174.5±110 in moderate, 114±70 in the severe group). But, the statistical difference has only been found in comparison to controls (773±356, p<0.01; Fig 1f). The mean percentage of CD16 on neutrophils from patients with COVID-19 was; 96.9%±2.8 in mild; 94%±11 in the moderate, and 96%±5 in the severe group of patients. No difference has been found between the groups even in comparison to controls (96%±5.5; p>0.05). MFI values for CD16 expression were 571±272 in the mild group, 385±210 in the moderate group and 536±248 in the severe group versus 1093±454. No difference was found among the groups. However, all was low in comparison to controls (p<0.01; Fig 1g, h). Therefore, we demonstrated that neutrophils of all patients with COVID-19 were phenotypically mature. The tendency to shed surface CD10 and CD16 could be a hallmark for their activation status.

### Full competency in ROS production and phagocytosis of neutrophils and monocytes in patients with COVID-19

The oxidative burst (OB) of neutrophils and monocytes is measured as a percentage value, as a result of the conversion of nonfluorescent dihydrorhodamine (DHR123) to fluorescent rhodamine 123 oxidized by the reactive oxygen species (ROS) produced in the stimulated cells. Percentage of OB in both monocytes and neutrophils of Covid-19 patients was greater than 95% similar to the control group indicating that no defect in ROS production of monocytes and neutrophils of COVID-19 patients (Fig 2). Similar results were obtained from the phagocytosis of fluorescent-labeled E.coli (>%90 in both monocytes and neutrophils). In addition to percentage results, the folds increase was calculated by dividing the MFI values from each stimulated sample to samples’ own initial MFIs. The folds increase in ROS production in the OB experiments was not different between the patients and controls (2.3±0.2 fold for neutrophil OB; 2.1±0.1 fold for monocyte OB in both groups). But, the folds increase in phagocytosis has found to be slighty low in patients with COVID-19 (2.07±0.1fold in the patient group vs 2.3 ±0.17 fold in the controls for neutrophils; 2.09±0.08 fold in the patient group vs 2.3±0.08 fold in the controls for monocytes; p=0.02). This decrease may be due to the fact that neutrophils are busy due to viral infections and its consequences such as clearance of necrotic and apoptotic cells. Thus, neutrophils and monocytes of patients with COVID-19 were shown to be fully competent in ROS production.

**FIGURE 2.**
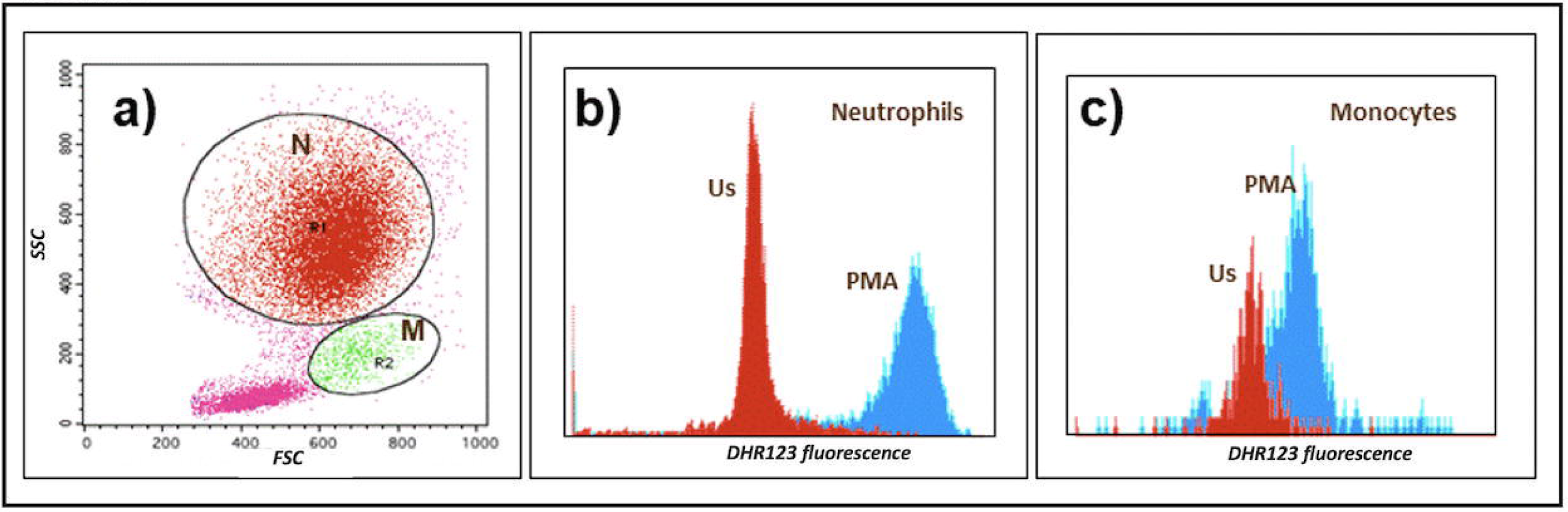
A representative figure for neutrophil and monocyte oxidative burst in a patient with COVID-19. **a)** Neutrophils (N) and monocytes (M) were gated on forward vs side scatter plot **b)** Neutrophil oxidative burst as DHR123 fluorescence (Us: unstimulated; PMA stimulated) **c)** Monocyte oxidative burst as DHR123 fluorescence (Us: unstimulated; PMA stimulated)

### Apoptosis of monocytes and neutrophil

No statistical significance has been found in early and late apoptosis of monocytes and neutrophils between the patients with COVID-19 and controls (data not shown).

### CD3^-^CD8^+^CD56^+^ NK like innate lymphoid cells

While evaluating CD3 vs CD8 plots during lymphocyte subset analysis, it has been noticed that there was an increase in CD3^-^ CD8^+^ quadrant. Then, it was shown that this population was CD56^+^ (Figure 3a, b). The percentages of CD3^-^ CD56^+^cells gated on CD8^+^ cells were calculated in all patient groups and controls. This was 18.2% ±10 in the control group, 33.8%±14 in the mild; 33.8%±22 in the moderate and, 41%±19 in the severe group (Fig 3c). Thus, we demonstrated that the percentage of CD3^-^CD8^+^CD56^+^ NK like innate lymphoid cells were significantly high in patients with COVID-19 (p<0.001).

**FIGURE 3.**
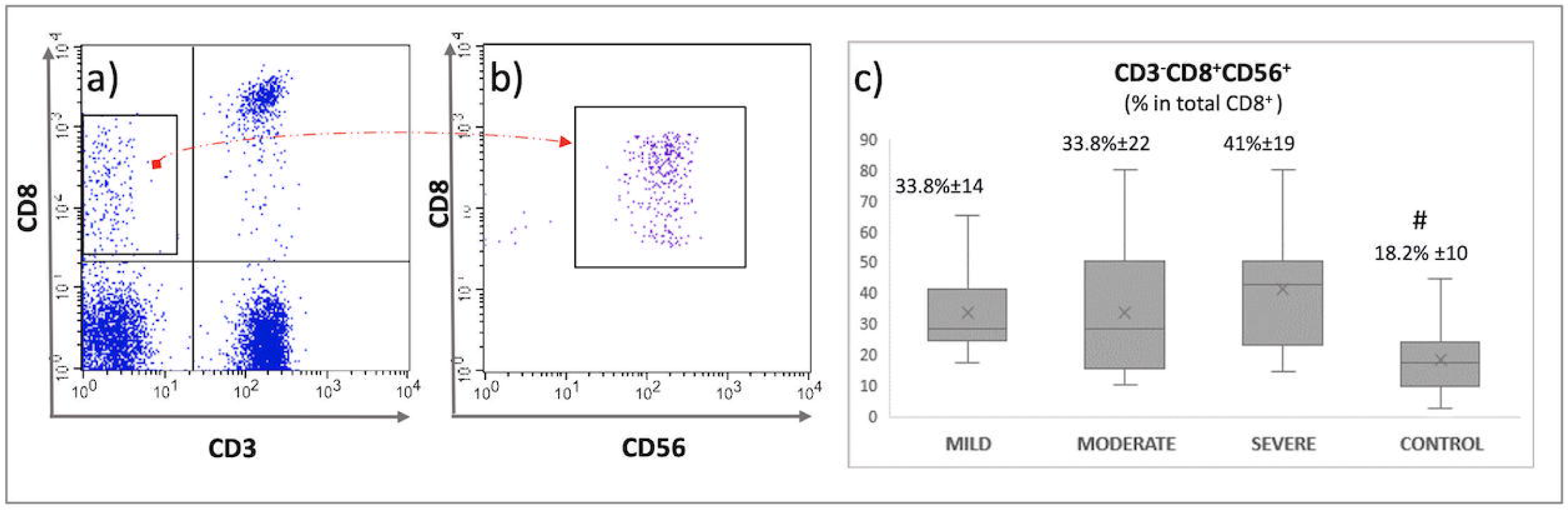
Increased CD3-CD8+CD56+ NK like innate lymphoid cells in COVID-19: **a)** A representative CD3 vs CD8 plot from lymphocytes of a patients with COVID-19. CD3^-^CD8^+^ cells were gated. **b)** CD56 expression of CD3^-^CD8^+^ cells. **c)** The percentages of CD3^-^CD8^+^CD56^+^ cells were calculated on CD8 gated lymphocytes. This was significantly high in all patient groups with COVID-19 in comparison to control (#:p<0.01).

### NK cells

As bridging cells between innate and adaptive immune systems, the percentages and absolute numbers of NK cells (CD3^-^CD16^+^CD56^+^) were analyzed. Results for all three groups of patients with COVID-19 and controls were given in Table 4. Although there was an elevation tendency in NK cells in the severe group and, remarkably high percentage of NK cells was observed in some of the severe cases, no statistical difference has been found among the groups (Figure 4c). The absolute number of NK cells was found to be low in all groups of patients in comparison to healthy controls (p<0.03, Table 4) Thus, it was shown that the percentage of NK cells was in the normal range but, their absolute numbers in all groups of COVID-19 patients were as low as approximately 50% of controls.

**TABLE 4.**
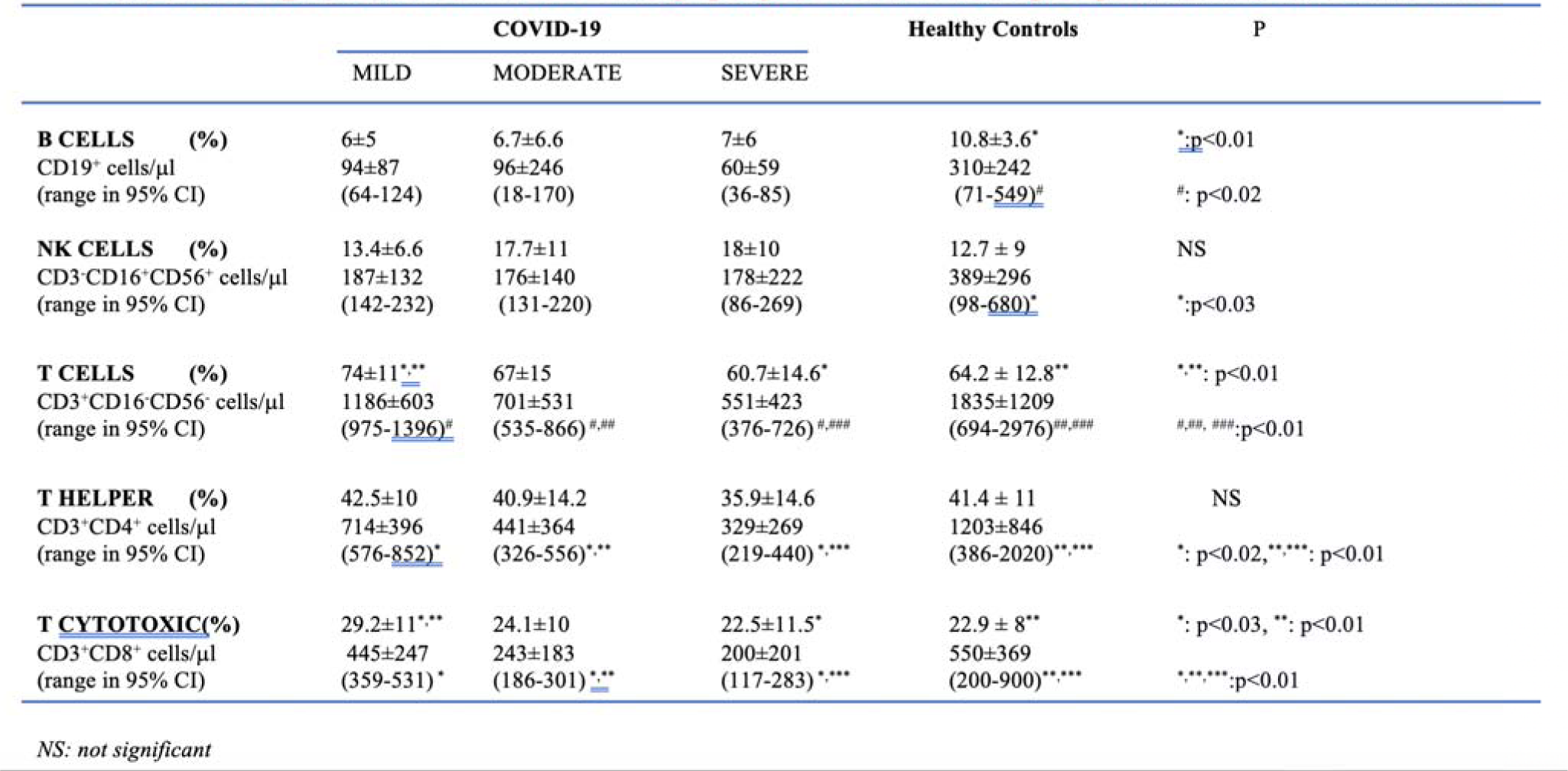
Percentages and absolute numbers of main lymphocyte subsets in various stages of patients with COVID-19 and controls.

**FIGURE 4.**
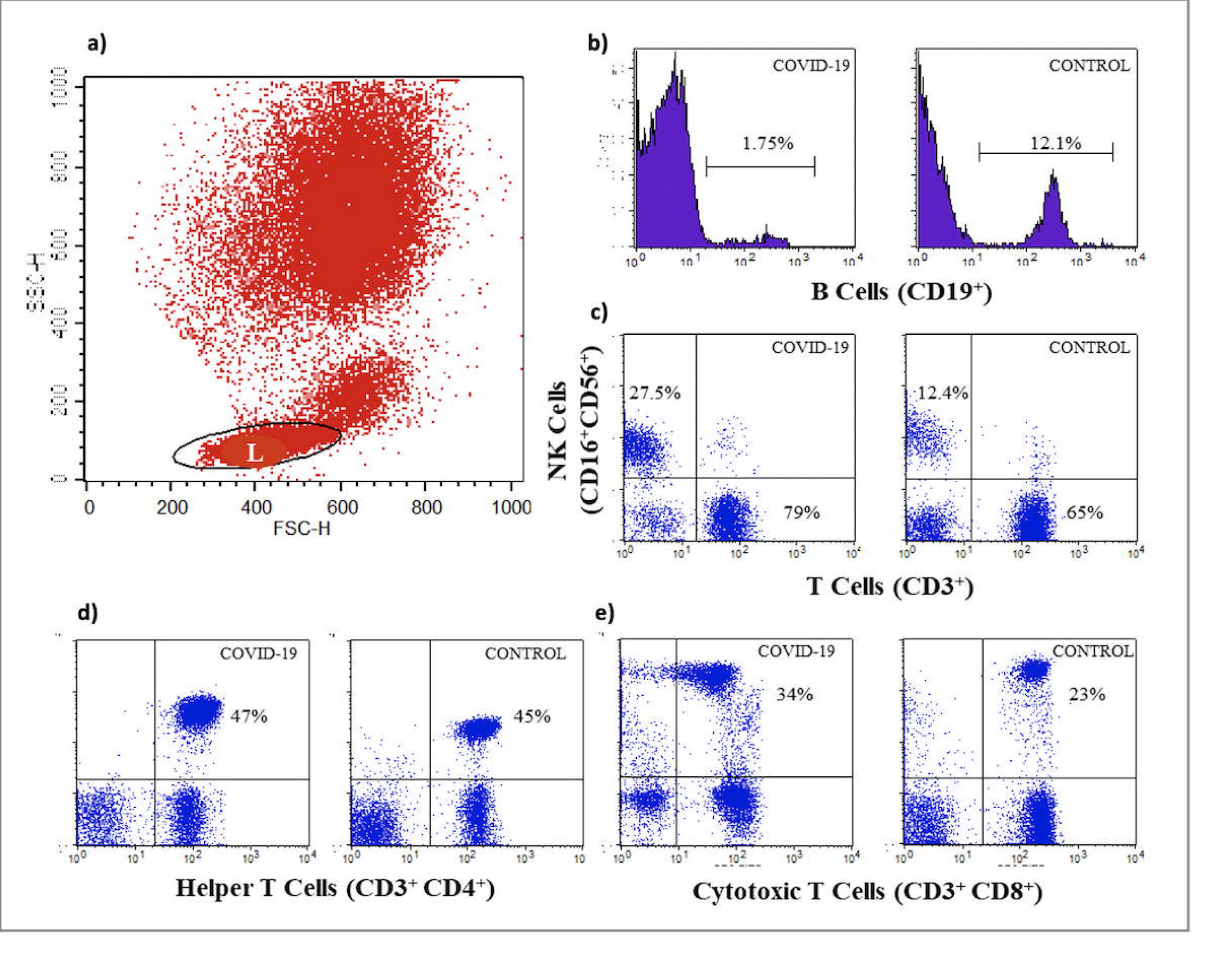
Representative flow cytometric plots for the main lymphocyte subsets in a patient with COVID-19 and a control. **a)** Lymphocytes (L) were gated on forward vs side scatter plot **b)** Histograms for CD19^+^ B cells. **c)** Plots demonstrating NK (CD3^-^CD16^+^CD56^+^) and T (CD3^+^CD16^-^CD56^-^) cells. **d)** Representative plots for T helper (CD3^+^CD4^+^) **e)** and T cytotoxic/suppressor cells (CD3^+^CD8^+^).

In the evaluation of phenotypic and functional features of the innate immune system in COVID-19 demonstrated that the presence of fully functional and active monocytes and neutrophils, and the normal or an increased percentage of bridging cells such as NK and NK like innate immune cells, respectively. However, the decreased absolute number of NK cells was not accorded with their percentage values.

### Phenotypic features of adaptive immune system cells in COVID-19

A representative figure for flow cytometric evaluation of lymphocyte subsets was shown in Figure 4. All percentages and the absolute numbers of lymphocyte subsets was given in Table 4.

### The percentage and the absolute number of B lymphocytes were extremely low in all groups of COVID-19 patients

The percentage of B lymphocytes evaluated by surface CD19 was low in all COVID-19 patients compared to control (p <0.01, Figure 4b, Table 4). There was no difference among patient groups. Furthermore, when the absolute B lymphocyte counts were examined, the number of B lymphocytes was found to be remarkably decreased in patients with COVID-19 (p<0.02, Table 4). In the severe group, the number of B lymphocytes decreased up to approximately one-fifth of the control. CD19 and CD138 were stained in a few patients with the thought that the reason for this extreme reduction may be the result of the differentiation of B lymphocytes into plasma cells. However, no CD138 positive cells were found in the periphery.

### The absolute number of T cells gradually decreased according to the severity of the disease

A significant increase in the percentages of T cells (CD3^+^CD16^-^CD56^-^) was found in the mild group of patients with Covid-19 in comparison to the severe group and healthy control, as well (p<0.01, Table 4, Figure 4c). The absolute number of T cells was in the normal range in only the mild group of patients whereas this was found to be decreased in both moderate and severe groups in comparison to the mild group of patients and controls (p<0.01, Table 4).

### No difference in the percentage of T helper cells but its number decreased in the moderate and severe COVID-19 groups

The percentage of CD3^+^CD4^+^ T cells was not different among the groups (Table 4, Figure 4d). The absolute number of CD3^+^CD4^+^ T cells was in the normal range in the mild group. But, this was significantly reduced in moderate and severe disease courses in comparison to controls and also mild disease course (p<0.01; p<0.02 respectively, Table 4).

### A normal number of T cytotoxic/suppressor cells with an increased percentage in the mild group decreased with the disease severity

The percentage of CD3^+^CD8^+^ cytotoxic/suppressor T cells was found to be elevated in the mild group in comparison to severe patients (p<0.03) and controls, as well (p<0.01; Table 4, Figure 4e). Thus, it was shown that the reason for the increase in the percentage of CD3^+^T cells in the mild group was due to the increase of CD8 ^+^ T cells. The absolute number of CD3^+^CD8^+^ T cells was in the normal range in the mild group. But, this was significantly decreased in the moderate and severe groups in comparison to the mild group and healthy controls (p<0.01, Table 4). When the CD4 ^+^CD8^+^ double-positive cells and the ratio of CD4/CD8 were examined among all the groups, no difference was detected.

### Spontaneously activated CD4^+^ and CD8^+^ T cells in COVID-19

We studied CD69 expression on T lymphocytes without any stimulation to measure spontaneous activation. In the severe group of patients with COVID-19, a significant increase has been found in the expression of CD69 on CD4^+^ (3.7% ±4.9 in the mild; 3.6% ±4 in the moderate; 9.4% ±6.8 in the severe group; p<0.01 with severe group; Fig 5a, b) and on CD8^+^ cells (13.3% ±12.4; 30% ±35; 45% ±34.5 in the mild, moderate and severe groups respectively (p=0.01 between mild and severe group; Fig 5c, d).

**FIGURE 5.**
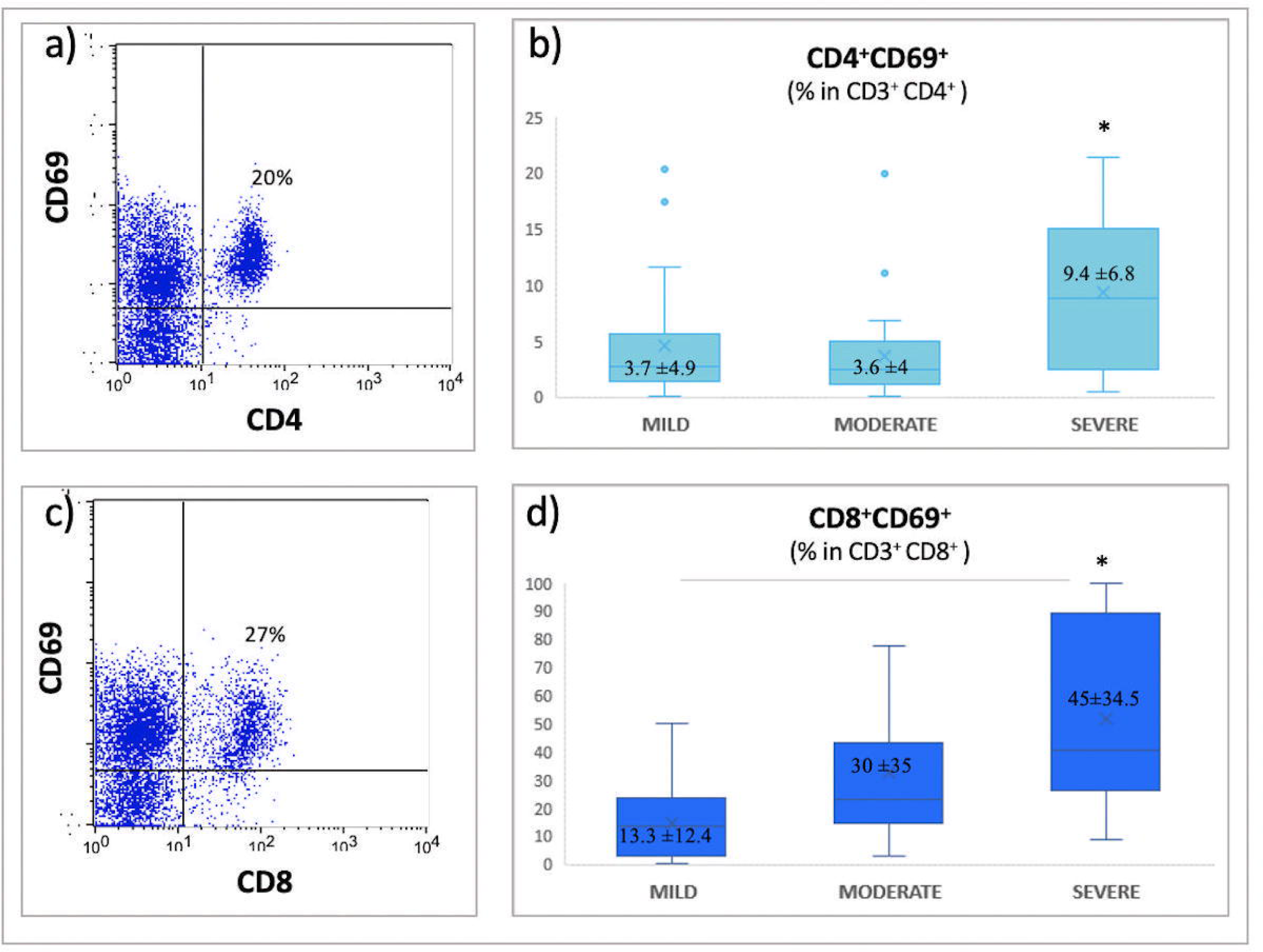
Spontaneous CD69 expression on CD4+ and CD8+ T cells in patients with COVID-19. **a, c)** Representative CD69 vs CD4 and CD8 plots demonstrating spontaneous CD69 expression on lymphocytes from a patient with COVID-19 **b)** The ratio of CD69 on CD4+ T cells was significantly high in the severe patients (9.4% ±6.8) in comparison to the mild (3.7% ±4.9) and moderate patients (3.6% ±4; *: p<0.01). **c)** The ratio of CD8+CD69+ T cells was significantly increased in the severe group (45% ±34.5) in comparison to the mild group (13.3% ±12.4; *: p=0.01)

### An increase in NK like T cells (NKT; CD3^+^CD56^+^) in patients with COVID-19

Although CD3 positivity is accepted as a marker of adaptive T cells, CD3^+^CD56^+^ T cells represent a heterogeneous group of innate cells with their expression of restricted range of T cell receptor (TCR) that play roles in the immune response to tumors and pathogens(26, 27). In our study, the percentage of CD3^+^CD56^+^ population was found to be high in COVID-19 patients compared to controls (6.7%±4.3 vs 1.9%±2; p<0.01, Figure 6) No difference was found among patients.

**FIGURE 6.**
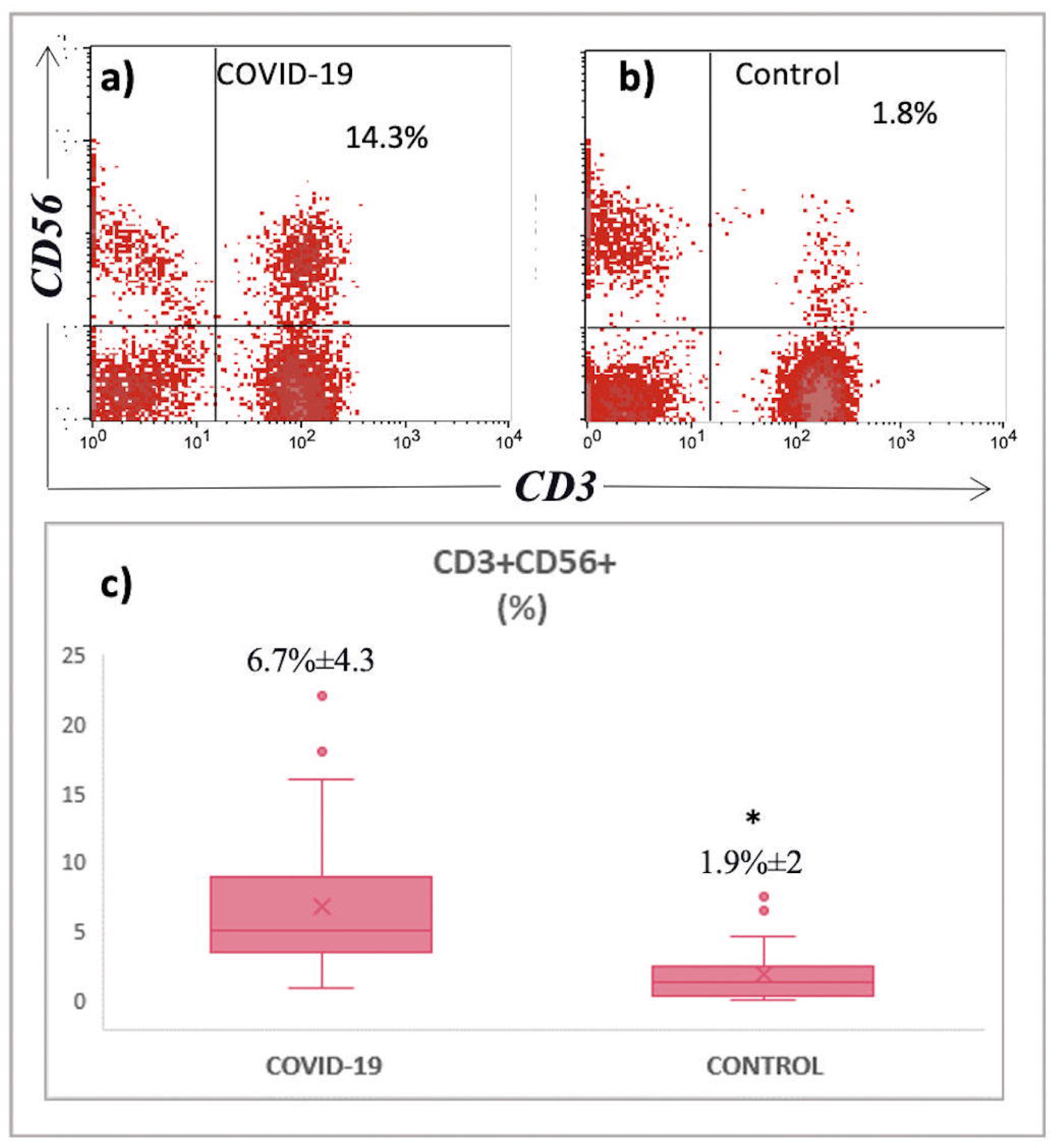
CD3^+^CD56^+^ NK like T cells were high in COVID-19. **a, b)** Representative CD3 vs CD56 plots from a patient with COVID-19 and a control. **c)** *: p<0.01: The percentage of CD3^+^CD56^+^ T cells was significantly high in patients with COVID-19.

Interestingly, we found that CD25 expression of T cells was very low (CD25 <15% on T cells) in all COVID-19 patients (data not shown).

### Suppressive lymphocte subset in COVID-19

It has been known that CD3^+^CD4^-^CD8^-^ double-negative T cells are suppressive T cells (28) The percentage of CD3^+^CD4^-^CD8^-^ was 3.6%±3.4 in the mild group; 4.5%±3.6 in the moderate group;5.4%±4.5 in the severe group and 2.1%±1 in controls. No difference has been found between controls and mild group, however a significant difference was observed between controls and both moderate and severe groups (p<0.01; Fig 7a, b, c).

**FIGURE 7.**
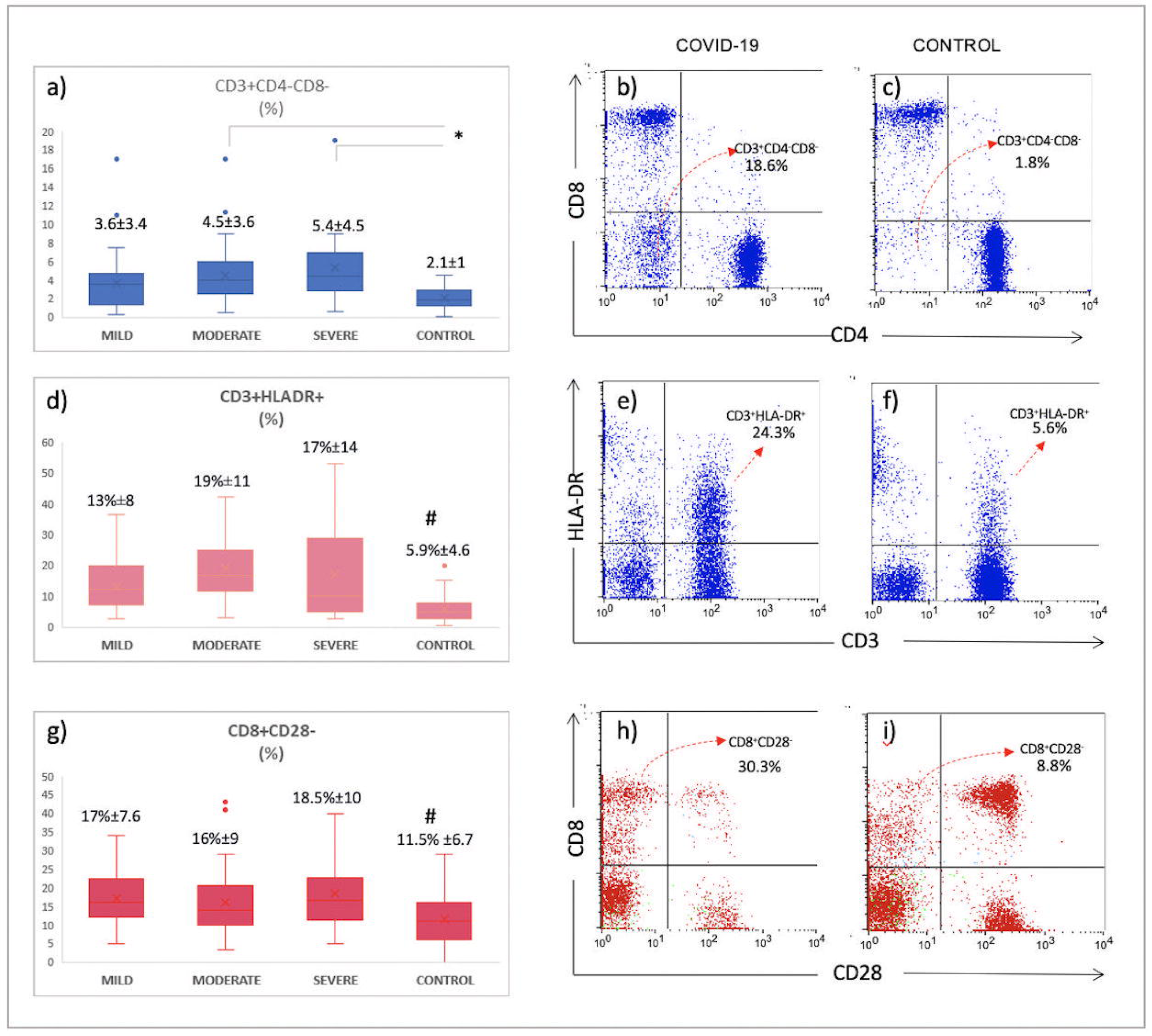
Suppressive T lymphocyte subsets increased in patients with COVID-19. **a)** The ratio of double-negative T cells (CD3^+^CD4^-^CD8^-^) was significantly increased in patients with COVID-19 with moderate or severe courses in comparison to the control group (*: p<0.01). **b)** Representative CD4 vs CD8 plots gated on CD3^+^ T cells in a patient with COVID-19 and **c)** healthy control. Lower left quadrant indicates CD3^+^CD4^-^CD8^-^ cells **d)** #: p<0.01: HLA-DR expressing T cells were increased in patients with COVID-19. **e, f)** CD3 vs HLA-DR plots from a patient with COVID-19 and a control. **g)** #: p<0.03: The percentage of CD8^+^CD28^-^ suppressive phenotype was prominent in COVID-19 cases. **h, i)** CD8 vs CD28 plots in patient with COVID-19 and a control. Upper left quadrant indicates CD28 negative CD8^+^ cells.

As an activation related suppression marker, HLA-DR on T lymphocytes was examined. **CD3**^+^ **HLA-DR**^**+**^ cells was elevated in all Covid-19 patients in comparison to controls in general (16%±11 vs 5.9%±4.6 respectively; p<0.01) This was 13%±8 in the mild; 19%±11 in the moderate and, 17%±14 in the severe group. (p<0.01; Fig 7d, e, f). No difference has been found among patients’ groups.

**CD8**^**+**^**CD28**^**-**^ T cells are a subset of regulatory T cells which has suppressive effect on CD4^+^ T cells (29, 30). In our study, the percentage of **CD8**^**+**^**CD28**^**-**^, suppressor T cells were found to be significantly increased in all groups of COVID-19 patients [17% ±8.9 (17%±7.6 in the mild, 16%±9 in the moderate 18.5%±10 in the severe group) vs 11.5% ±6.7 in the control group; p<0.03; Figure 7g, h, i)

### Lymphocyte apoptosis in the periphery depends on spontaneous activation of caspase-3 in patients with Covid-19

A special attention was focused on the lymphocyte population. Lymphocyte early apoptosis/activation rate (PhiPhilux only) following PMA stimulation was 36%±14.8 in patients with Covid-19 while that was 16.6%±6.2 in the control group (p=0.02; Fig 8a). Lymphocyte late apoptosis (PhiPhulux+PI stained cells) rate was 19.8%±11 in patients’ group; 9.6%±7.7 in the control group. Although Covid-19 patients showed higher late apoptosis on average, this difference did not reach statistical significance (Fig 8a). There was no difference between the two groups in the measurement of cell death, which was evaluated with PI stained only cells (20.5%±8.5 vs 16.8%±15.6; p>0.05). However, unlike controls, we observed that the relative number of PMA-induced lymphocytes in Covid-19 patients decreased significantly compared to their initial samples indicating spontaneous cell death. However, due to the small number of samples, we did not perform statistical analysis for this.

**FIGURE 8.**
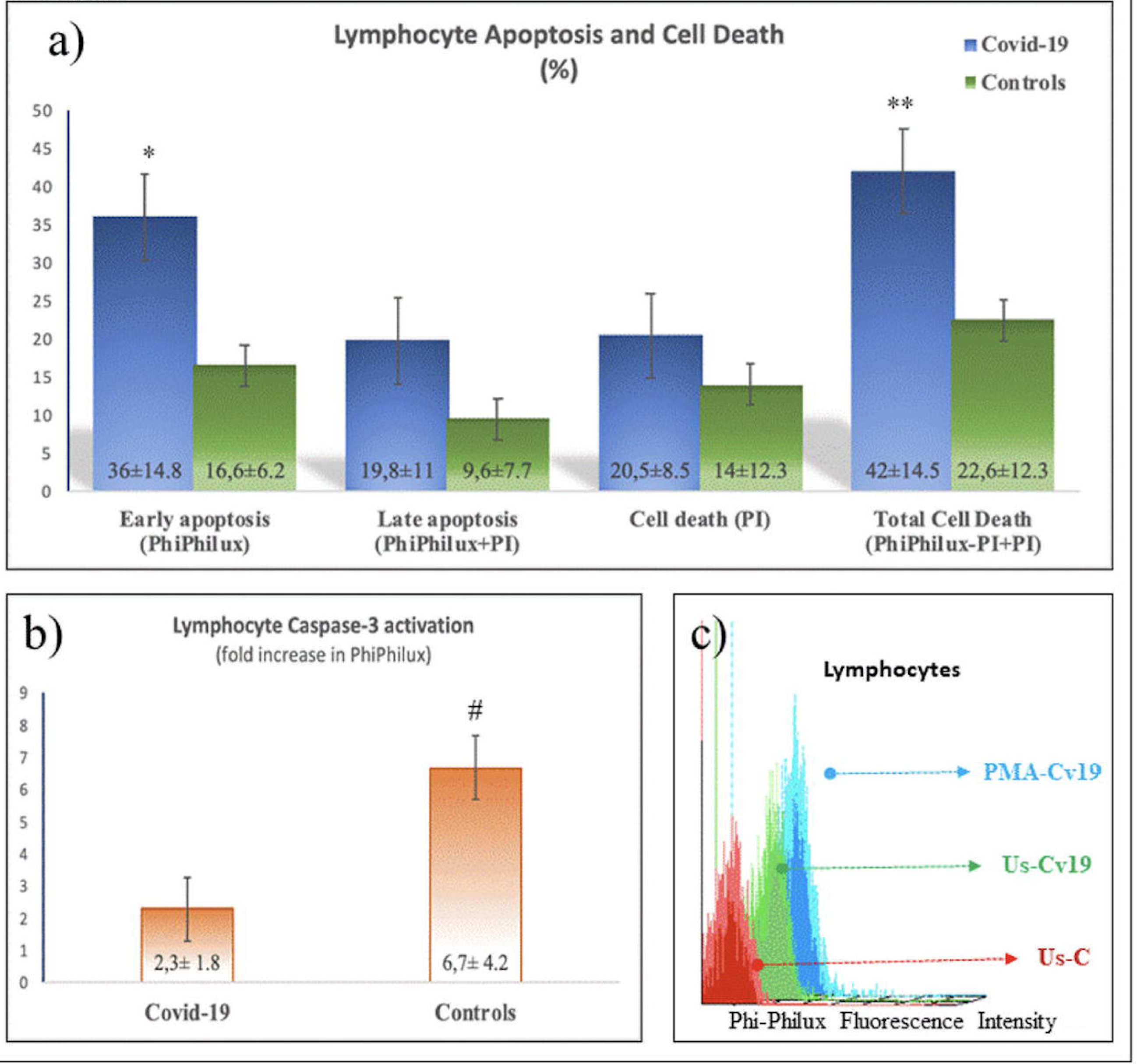
Lymphocyte early, late apoptosis, and cell death. a) Early apoptosis measured by the percentage of Phiphilux fluorescence was high in patients with Covid-19 (*: p=0.02). Total cell death as a sum of Phiphilux-PI-positive and PI only positive cells was elevated in patients with Covid-19 in comparison to controls (**: p<0.05). b) Fold increase in PhiPhilux fluorescence as an indicator of caspase-3 activation in lymphocytes was low in Covid-19 patients (#: p<0.01). c) A representative histogram showing baseline caspase 3 activations (PhiPhilux fluorescence intensity) in lymphocytes from a healthy control (UsC: unstimulated control), a Covid-19 patient (UsCv19: unstimulated Covid-19) and same Covid-19 patient following PMA stimulation (PMA-Cv19).

Total cell death was calculated by summing percentages of late apoptotic cells (PhiPhilux+PI) and cell death (PI only). This was significantly high in lymphocytes of Covid-19 patients (42%±14.5 vs 22.6%±12.3 in controls; p<0.05; Fig 8a). In addition to percentage increases, folds increases have been calculated by dividing PMA stimulated MFI value to the MFI value of the unstimulated sample for each sample. Interestingly, COVID-19 patients showed significantly lower fold increase than controls (2.3 ± 1.8 in Covid-19 patients, 6.7 ± 4.2 in controls; p <0.01; Fig 8b). It was an intriguing question of how come they have significantly low stimulation response in folds increase assessments while the percentage of lymphocyte early apoptosis following PMA stimulation was significantly higher in COVID-19 patients. To find an answer, we compared initial activation levels in unstimulated samples from both control and patient groups. In patients with COVID-19, initial fluorochrome conversion value for caspase-3 was significantly higher that indicated spontaneous activation of caspase-3 (17%±8 versus 6.3%±4.6; p=0.01; Fig 8c). Therefore, we thought that spontaneous caspase 3 activation could make COVID-19 lymphocytes more fragile to cell death. Thus, one of the causes of lymphopenia in COVID-19 patients might be elevated basal caspase-3 activation of peripheral lymphocytes for an unknown reason.

## DISCUSSION

The success for the immune system is a complex process that includes of innate at first followed by adaptive immune system activation in which controls innate system not to harm own cells with sustained reactions while producing specific, focused responses for eradicating the microorganism in the shortest time. At the same time it keeps memory cells while highly activated self cells are destroyed to avoid prolonged activation. Therefore, proper immune response against microorganisms including viruses depends on a fine tuned balance between innate and adaptive immune systems (11, 12, 31).

As a part of innate immune system cells, monocytes, as antigen-presenting cells, constitutively express Major Histocompatibility Complex (MHC) Antigen Class II, HLA-DR. This expression is a requirement for the communication between innate and adaptive immune systems. The peptides that were processed from microorganisms are presented by antigen-presenting cells to the T cells in the context of MHC molecules to activate adaptive T lymphocytes. The decrease in the expression of HLA-DR on monocytes is generally accepted as a marker of immune paralysis (20). In our study, we found decreased expression of HLA-DR on monocytes in all clinical courses of the patients with COVID-19 in comparison to healthy control. However, since we do not have a diseased control group, we did not have an idea of monocyte HLA-DR expression in viral diseases. A recent study showed that COVID-19 patients exhibited a less pronounced decrease in HLA-DR expression on monocytes in comparison to bacterial septic shock patients (13). On the other hand, CD16^+^ monocytes are a well-defined group known as intermediate and/or non-classical monocytes. They constitute 2-8% and 2-11% respectively of circulating monocytes under normal conditions (43). While CD16-monocytes secrete high levels of IL-6, CD16+ monocytes release high levels of IL-1β and TNF and are considered as pro-inflammatory monocytes (6). In our group of patients, we demonstrated more than 50% of monocytes of COVID-19 patients were CD16+ proinflammatory monocytes indicating that monocytes of COVID-19 patients actively contributed to the inflammatory process.

Neutrophils have critical roles in acute inflammation. Their functions are not limited to phagocytosis and oxidative burst, but also generates neutrophil extracellular traps (NETs) and modulate adaptive and innate immune responses (32, 33) Various neutrophil subgroups has also been defined according to their surface molecule profiles (34). One of the neutrophil surface molecules, CD10 (CALLA), is a neutral endopeptidase and cuts inflammatory peptides such as substance-P, met-enkephalin, fMLP. Those peptides play role in acute inflammation (35). Mature neutrophils are CD10 positive, immature ones are CD10 negative. Each has different effects on T lymphocytes. CD10 positive neutrophils inhibit interferon-γ release and proliferation of T lymphocytes, while CD10 negatives help T cell survival (36). CD16 is Fc gamma receptor III and is a necessary molecule for neutrophil phagocytosis (37). Following activation and apoptosis, neutrophil CD16 is cut by ADAM17 metalloprotease and it sheds from membrane(38). Thus, CD16 shedding is a marker for neutrophil activation and apoptosis. In our group of patients, CD10 and CD16 positive mature neutrophils were slightly decreased in the severe group. This reduction can be evaluated as leaving their place to immature neutrophils, in line with the severity of inflammation. Our finding that CD16 expression is not different from controls as a percentage can be considered as an indication that COVID-19 disease does not cause neutrophil apoptosis and neutrophils remain competent. Supporting this hypothesis, no defect in ROS production of neutrophils and monocytes has been found. A slight decrease in phagocytosis as MFI may indicate that neutrophils and monocytes of the patients with COVID-19 are busy. Our finding that early, late apoptosis and cell death in monocytes, especially in neutrophils, are not different from controls. Together with our all data on the innate cells, we consider that COVID-19 does not target the monocytes and neutrophils of innate immune system cells.

The percentage of NK cells was found to be normal even in the severe group of patients in contrast to the previous study (24, 25). A decrease was observed in the number of NK cells, but this reduction was not as excessive as in B cells. Based on the knowledge that SARS-CoV affects interferon pathways, NK functions may be considered to be decreased, but we could not perform functional studies for NK cells (39, 40). According to a previous study, NK functions measured by IFN-gamma release were found to be within normal limits (25). Future studies on direct cytotoxicity or antibody-mediated cytotoxicity of NK cells will help us to interpret this issue better. We identified a significant increase in CD3^-^CD8^+^CD56^+^ innate lymphoid cells and CD3^+^CD56^+^ NK like innate T cells in all COVID-19 patients. CD3^-^CD8^+^CD56^+^, innate lymphoid cells similar to NK cells are potent cytotoxic and have strong capacity to release IFN-γ (41–44). We suggest that the increase of CD3^-^CD8^+^CD56^+^ cells with their possible capacities of both cytotoxicity and releasing IFN-γ might demonstrate the cytotoxic/killer cells that try to respond rapidly to SARS-CoV-2 in COVID-19 patients. Increased expression of CD56 which is an adhesion molecule, may provide a privilege to these CD8^+^ cells to rapidly navigate between the periphery and the inflammation. It might be considered that the decreased number of NK cells could be compensated by increased innate lymphoid cells. The fact that we detected a innate NK, T cell dominant immune profile maybe because we have a trained innate immune system as a result of having a national compulsory vaccination program including BCG and measles vaccines. Our limitation was that we could not have measured cytokine release from isolated cell populations from patients. This issue requires further functional studies.

In our group of patients, we have found that the absolute number of B lymphocytes was extremely reduced. Obviously, we should not expect proper immunoglobulin secretion from plasma cells in COVID-19 with that low number of B lymphocytes. It has already been shown that there was no peripheral B cell memory in Severe Acute Respiratory syndrom in six years follow-up (17, 45). In our study, we considered that this decrease might have been related to plasma cell differentiation. However, we demonstrated that no plasma cells have found in the periphery. A second reason might be the migration of B lymphocytes to the inflammation site. But, histopathological studies did not support this hypothesis. In addition, there is no finding such as lymphadenopathy. Furthermore, it has been shown that there are no lymphocytes in lymph node and spleen (46) Another possibility is that, as a member of the adaptive immune system, B lymphocytes may be one of the targets that directly excluded by SARS-CoV-2. As a part of imbalanced immune behavior, imbalanced cytokine release caused by SARS-CoV-2 may not permit proper involvement of B lymphocytes. More studies on B lymphocytes are require.

The significant increase in the percentages of CD3^+^ and CD8^+^ T lymphocytes with their normal ranged absolute numbers in the mild patients can be considered as a sign of a strong immune response at the beginning. In the mild group, while the absolute number of CD4^+^ T cells are still within normal limits, this decrease in both NK and B cell populations suggests that a careful phenotypic follow-up can be used to predict disease progression from the beginning. While the decrease in the absolute numbers of NK and B cell remained the same throughout the disease progression in all patient groups, a sharper decrease in CD4 and CD8 T lymphocytes in the moderate group is noteworthy, and this decline continues as the disease becomes severe. Even that decrease in the number of CD4^+^ T cells in the severe group approaches the decrease in B lymphocytes.

One of the regulatory T cell populations, CD3^+^CD4^-^CD8^-^T cells constitute 1% of peripheral T cells (47). It has been previously shown that they have a strong suppressive effect on CD4^+^ and CD8^+^ cells, (28). Our finding that increased CD3^+^CD4^-^CD8 T cells especially in moderate and severe groups suggests that the activation control of the adaptive immune system is strong in COVID-19 even if it is not required. HLA-DR expression on T lymphocyte has long been known to down-regulate T cells that have already activated and to represent an important homeostatic regulatory mechanism(48). CD8^+^CD28^-^ T cells represent another group of regulatory T cells that inhibit CD4^+^ T cell proliferation(29, 30). In our study, the increase of CD3^+^ HLA-DR^+^ and CD8^+^ CD28^-^ T lymphocytes also indicate the presence of a tight adaptive immune down-regulation in COVID-19.

Our finding that early apoptosis or apoptotic activation in lymphocytes, which we measured with the conversion of a non-fluorescent substrate into fluorescence when is cut by active Caspase 3, indicates that the virus leads a process that initiates caspase 3 activation in lymphocytes although we do not know the interacting molecules yet. Indeed, a recent sophisticated study demonstrated that 332 human SARS-Cov-2 interacting proteins have been defined (31). Our finding that folds increase upon stimulus in caspase 3 activation was low in the patient group in contrast to controls suggests that COVID-19 patients have pre-activated/tired and busy lymphocytes. Caspase-3, which was already active at the basal level, cannot be activated further by stimulation. But even with this tired activation level, it is able to double the total lymphocyte death rate compared to the controls. This data can be interpreted as an indication that SARS-Cov-2 is destructive for peripheral lymphocytes. Already, an increase in LDH in patients is a sign of active destruction.

In conclusion, this study, in which we examine partially the phenotypic and functional characteristics of the innate and adaptive immune system cells, shows that the innate immune system functions in COVID-19 are complete and competent, and the remaining a low number of lymphocytes is half-dead trying to respond to viral infection with a suppressive profile in COVID-19. It supports our hypothesis that innate-adaptive immune system communication is impaired in COVID-19. In this context, it will be very important to focus on additional functional studies related to apoptotic targets in explaining immune pathogenesis.

## Data Availability

All data are available upon request from the first author.

## CONFLICT OF INTERESTS

No conflict of interest for none of the authors

## AUTHOR CONTRIBUTIONS

E.E.D: conceptualized and designed the study, performed partly lab experiments, analyzed the data, wrote the manuscript, drafted the manuscript, and gave approval of the final version to be submitted. S.A., U.S., D.B., İ.O., N.A., S.G. collected blood samples, collected clinical information, reviewed the manuscript, and gave approval of the final version to be submitted. R.Y. performed lab experiments, analyzed the data, reviewed the manuscript, and gave approval of the final version to be submitted. A.Y. contributed designation of the study, reviewed the manuscript, and gave approval of the final version to be submitted.

## FUNDING

The study was funded in part personally by Dr. Ekşioğlu-Demiralp and, in part by Memorial Health Group.

## ACKNOWLEDGMENTS

A special thanks to Berna Altun, Damla Kılıç, Gamze Ensar and Gözde Akçin from Memorial Tissue Typing and Immunology Laboratory and Melek Yildiz from Department of Infectious Disease for their valuable contributions to the lab works and collecting patient information.

## Notes

### Competing Interest Statement

The authors have declared no competing interest.

### Author Declarations

Marmara University Ethics Committee (no:08.05.2020/09.2020.541

